# Handling Missing Data in Participants with a Baseline but No Post-baseline Data

**DOI:** 10.1101/2025.03.26.25324679

**Authors:** Craig Mallinckrodt, Ilya Lipkovich, Samuel Dickson, Suzanne Hendrix, Geert Molenberghs

## Abstract

Clinical trial participants who are randomized to treatment and have a baseline value but no post-baseline data pose a unique challenge. These patients need to be included to preserve randomization. However, there is no information about the outcome or the intercurrent events that led to missing data. Composite and hypothetical strategies can accommodate participants with no post-baseline data. The present study investigated common methods based on hypothetical strategies in example data sets and in a simulation study. Because there is no information about the treatment effect when there is no post-baseline data, it is not surprising that various models yielded similar results. In simulated data, treatment contrasts were not biased when the reason for missing all post-baseline data was random, treatment related, or outcome related. Bias occurred only when missingness was treatment and outcome related, and arose from a missing not at random mechanism. The bias was not large. The maximum Type I error rate across all scenarios and methods was 6.4%. Imputing no change for missing data at the first postbaseline visit and assuming missing at random for all other missing values yielded a maximum Type I error rate of 5.1%. Given the idiosyncratic nature of clinical trials, no universally best analytic approach exists for dealing with participants that have a baseline but no post-baseline data. Analysts can choose among these methods to provide an approach tailored to the situation at hand.

## 1. Introduction

Consider clinical trial participants who are randomized to treatment and have a baseline value for the primary endpoint, but all post-baseline data are missing. This missing data problem can be dealt with using the ICH E9 (R1)^1^ estimands framework, as is done for other missing data scenarios^2^. However, the challenge in handling participants with no post-baseline data is that there is no information about either the outcome of interest or the intercurrent events (ICEs) that led to missing data. How do we account for an ICE when we do not know what it was?

This scenario has received increased attention from regulators recently. Sponsors now commonly receive feedback that FDA no longer accepts a modified intention-to-treat (mITT) approach that includes only participants with a baseline and at least one post-baseline observation. Instead, the primary analyses should include all randomized participants (ITT) and specify how missing data will be handled for participants with no post-baseline observations.

It is worth considering why mITT is widely accepted long after the benefits of ITT became clear. Perhaps mITT retained popularity via justification that participants with no post-baseline data provide no information about the treatment effect, therefore baseline data from these participants could be excluded without consequence. However, the reason the data are missing could be related to treatment and / or outcome; that is, the data may not be missing completely at random. Hence, participants with no post-baseline data cannot be ignored because ignoring them might jeopardize randomization.

This paper discusses the problem of participants with no post-baseline data in the estimands framework and compares the empirical performance of various analyses relative to common estimands.

In the next section, analytic alternatives for dealing with participants that have a baseline but no post-baseline data are described and two examples are introduced and analyzed. Section 3 details a simulation study to assess the magnitude of bias from participants that have no post-baseline data using various missing data mechanisms and magnitude of correlations between baseline and post-baseline values. Section 4 discusses and puts into context the results from Sections 2 and 3. Sections 5 has concluding statements.

## 2. Analytic alternatives for participants with no post-baseline observations

### 2.1 Example one data

The first example is a contrived data set, for convenience and simplicity (Table 1a), that has 10 participants per arm, with 1 participant in each arm having a baseline but no post-baseline data. Visit-wise means for each group are summarized in Table 1b. The actual post-baseline observations are the same in the ITT and mITT dataset. However, the baseline values are not the same, and therefore the mean changes also differ.

**Table 1a.** Example One Data Set.

| Obs | Sub-<br>ject | Treat-<br>ment | Base-<br>line | Y1<sup>1</sup> | Y2 | Y3 |
| --- | --- | --- | --- | --- | --- | --- |
| 1 | 2 | 1 | 20 | 14 | 12 | 15 |
| 2 | 5 | 1 | 12 | 6 | 9 | 3 |
| 3 | 6 | 1 | 14 | 8 | 4 | 4 |
| 4 | 8 | 1 | 21 | 19 | 12 | 12 |
| 5 | 9 | 1 | 19 | 10 | 13 | 9 |
| 6 | 13 | 1 | 20 | 11 | 8 | 7 |
| 7 | 14 | 1 | 19 | 13 | 7 | 3 |
| 8 | 16 | 1 | 19 | 16 | 8 | 2 |
| 9 | 18 | 1 | 23 | 16 | 13 | 8 |
| 10 | 19 | 1 | 26 | . | . | . |
| 11 | 1 | 2 | 24 | 13 | 8 | 0 |
| 12 | 3 | 2 | 18 | 17 | 17 | 9 |
| 13 | 4 | 2 | 10 | 19 | 4 | 1 |
| 14 | 7 | 2 | 17 | 10 | 10 | 3 |
| 15 | 10 | 2 | 19 | 6 | 7 | 5 |
| 16 | 11 | 2 | 20 | 9 | 5 | 0 |
| 17 | 12 | 2 | 19 | 12 | 4 | 0 |
| 18 | 15 | 2 | 19 | 7 | 4 | 1 |
| 19 | 17 | 2 | 20 | 11 | 7 | 1 |
| 20 | 20 | 2 | 19 | . | . | . |

1. Y1, Y2, and Y3 are the observations at Times 1, 2, and 3, respectively.

**Table 1b.** Descriptive Statistics for the Example One Data Set.

| **ITT Data set** |  |  |  |  |  |
| --- | --- | --- | --- | --- | --- |
| Time | Means<sup>1</sup> |  | Mean |  | Difference |
|  | Group 1 | Group 2 | Group 1 | Group 2 |  |
| 0 | 19.30 | 18.50 |  |  | 0.80 |
| 1 | 12.55 | 11.55 | 6.75 | 6.95 | 1.00 |
| 2 | 9.55 | 7.33 | 9.75 | 11.17 | 2.22 |
| 3 | 7.00 | 2.22 | 12.30 | 16.28 | 4.78 |

| **mITT Data Set** |  |  |  |  |  |
| --- | --- | --- | --- | --- | --- |
| Time | Means<sup>1</sup> |  | Mean within Group Change |  | Mean Difference |
|  | Group 1 | Group 2 | Group 1 | Group 2 |  |
| 0 | 18.55 | 18.44 |  |  | 0.11 |
| 1 | 12.55 | 11.55 | 6.00 | 6.89 | 0.89 |
| 2 | 9.55 | 7.33 | 9.00 | 11.11 | 2.11 |
| 3 | 7.00 | 2.22 | 11.55 | 16.22 | 4.67 |

### 2.2. Example two data

The second example includes two nearly identically designed clinical trials in major depressive disorder.^3,4^ These studies have been used as example data sets in prior research on the handling of missing data.^5,6^ Each trial had four treatment arms, with two doses of an experimental medication (subsequently granted marketing authorizations in most major jurisdictions), an approved medication, and placebo. Assessments on the Hamilton 17-item rating scale for depression^7^ were taken at baseline (week 0) and weeks 1, 2, 4, 6, and 8 in each trial.

For the example analyses, the three active arms from each trial were pooled and 1/3 of those patients were randomly selected to create a single active arm to be compared to the placebo group.^5,6^ This was done to avoid marketing implications for the active drugs. These trials are referred to as the low and high dropout datasets.

Descriptive statistics for these two example data sets are summarized in Table 2. The percentages of patients with no post-baseline data were 2.4% in the low dropout data set and 6.1% in the high dropout dataset. The corresponding overall dropout rates were 10.2% and 39%, respectively. The design features thought to contribute to the difference in dropout rates between these two otherwise similar trials was that the low dropout dataset came from a study conducted in Eastern Europe that included a 6-month extension treatment period after the 8-week acute treatment phase, and used titration dosing. The high dropout data set came from a study conducted in the US that did not have the extension treatment period and used fixed dosing.^5,6^

**Table 2.**
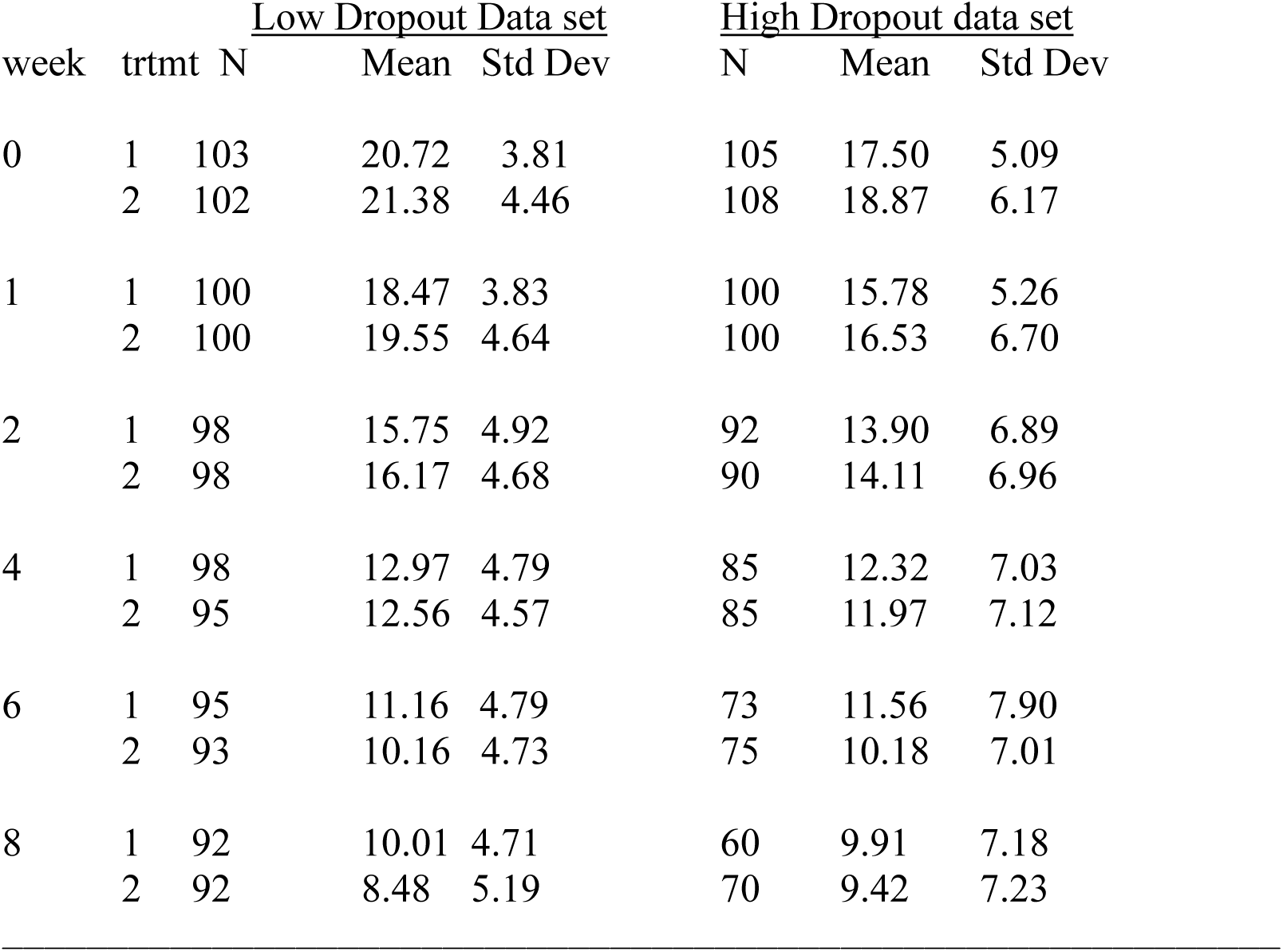
Descriptive Statistics for the Example Two Datasets

| week | trtmt | Low Dropout Data set |  |  | High Dropout data set |  |  |
| --- | --- | --- | --- | --- | --- | --- | --- |
|  |  | N | Mean | Std Dev | N | Mean | Std Dev |
| 0 | 1 | 103 | 20.72 | 3.81 | 105 | 17.50 | 5.09 |
|  | 2 | 102 | 21.38 | 4.46 | 108 | 18.87 | 6.17 |
| 1 | 1 | 100 | 18.47 | 3.83 | 100 | 15.78 | 5.26 |
|  | 2 | 100 | 19.55 | 4.64 | 100 | 16.53 | 6.70 |
| 2 | 1 | 98 | 15.75 | 4.92 | 92 | 13.90 | 6.89 |
|  | 2 | 98 | 16.17 | 4.68 | 90 | 14.11 | 6.96 |
| 4 | 1 | 98 | 12.97 | 4.79 | 85 | 12.32 | 7.03 |
|  | 2 | 95 | 12.56 | 4.57 | 85 | 11.97 | 7.12 |
| 6 | 1 | 95 | 11.16 | 4.79 | 73 | 11.56 | 7.90 |
|  | 2 | 93 | 10.16 | 4.73 | 75 | 10.18 | 7.01 |
| 8 | 1 | 92 | 10.01 | 4.71 | 60 | 9.91 | 7.18 |
|  | 2 | 92 | 8.48 | 5.19 | 70 | 9.42 | 7.23 |

### 2.2 Estimand considerations

When ITT-based analyses are needed, principal stratification strategies are not relevant.^8^ While on treatment (WoT) strategies are also not relevant because this approach also excludes participants with no post-baseline data. Treatment policy strategies ignore intercurrent events (ICEs), which are post randomization events. Hence, treatment policy strategies are not relevant to this scenario because there are no post-randomization events. Composite and hypothetical ICE strategies are both relevant to accounting for participants with no post-baseline data.

Composite strategies for dealing with ICEs that lead to missing data were in use long before defining estimands was common place.^9^ Composite strategies consider relevant ICEs to be an outcome, and combine ICE outcomes and observed outcomes into a single endpoint. Typically, the outcome ascribed by a composite strategy is a “bad” outcome reflecting a failed or ineffective treatment.

Historically, common composite strategies include non-responder imputation (NRI) for binary endpoints and baseline or last observation carried forward (BOCF, LOCF) for continuous endpoints. Modified approaches to NRI, BOCF, and LOCF (mNRI, mBOCF, mLOCF) can be applied when occurrence of an ICE does not always mean the treatment failed. In the modified versions a set of treatment-related reasons for treatment discontinuation, such as lack of efficacy or intolerability is pre-specified; NRI, BOCF, or LOCF are applied for these ICEs. However, discontinuations for reasons unrelated to study treatment, such as having relocated and no longer being able to attend study visits, are not relevant ICEs.^2,10^ NRI, BOCF, and LOCF have been criticized on a number of statistical grounds.^2,5,6,10,11^

Intuitively, BOCF and LOCF would not make sense for progressive diseases where no change is a treatment goal, or in situations such as example two in which there is an appreciable placebo response. For scenarios in which assuming no change for participants who discontinue is reasonable, more principled multiple imputation-based approaches to BOCF are available.^12^ One simple approach is to impute only the first post-baseline assessment using BOCF and leave the remaining assessments as missing, followed by application of an MAR-based analysis.

Non-parametric methods based on ranks or ‘scores’ of the observations instead of the actual values are readily applied in a composite strategy.^2,10^ ICEs are accounted for in these methods by ranking the events relative to actually observed outcomes according to the ICE severity, and the resultant rankings are analyzed via Wilcoxon signed-rank tests or Mann-Whitney tests.^13^ Rankings for ICEs leading to early study discontinuation could be based on the reasons for withdrawal^14^ or the time of withdrawal.^15^ For example, death may receive the worst rank, followed by discontinuation for lack of efficacy / adverse reactions, etc. Within the same category of withdrawal, early dropouts could get worse ranks than later dropouts.

The win ratio approach overcomes some of the difficulties in interpreting differences in ranks.^16^ Participants in the experimental and control treatments are paired in all possible combinations. Participants in the experimental treatment arm are labeled as winner (loser) if they have the more (less) favorable outcome. The population parameter estimated is the win ratio, which is the total number of winners divided by the total number of losers. Confidence intervals can be obtained using the bootstrap method. However, this method is not recommended when the number of ties is appreciable.^17^ A similar alternative is the win-odds method.^17^ An extensive review of these methods and other generalized pairwise comparison methods is available.^18^

An important consideration for any composite strategy when there is no post-baseline data is that there is no information about the outcome or the ICEs that led to dropout. Therefore, there is no data-based way to assess whether the outcome ascribed by the composite strategy is appropriate. Consider the example two data sets. Antidepressants are known to have slow onset of action. ^20^ Therefore, leaving the study before the first assessment is unlikely related to the longer-term efficacy of the drug. Perhaps participants left the study due to an adverse event, or perhaps they simply changed their mind and didn’t want to participate. Again, it can be hard to account for an intercurrent event when we don’t know what it was.

Hypothetical strategies are predicated on wanting to know what outcomes would have been under hypothetical conditions. Two hypothetical strategies that could be applied to participants with no post-baseline data are: 1) outcome trajectories that would have been observed in the absence of the ICEs; and, 2) outcome trajectories based on the assumption that if participants do not adhere to the medication, they will not benefit from it.^5^ The latter approach can be used as either a sensitivity assessment for approach 1, or as a more principled alternative to BOCF and LOCF for ascribing no benefit to patients with relevant ICEs by using placebo group outcomes to define no benefit. ^6,21–25^

Approach 1 assumes that missing data arise from a missing at random (MAR) mechanism; that is, conditional upon the covariates in the analysis and the observed outcomes of the variable being analyzed, the probability of missingness does not depend on the unobserved outcomes of the variable being analyzed.^5^ Approach 2 assumes that missing data arise from a missing not at random (MNAR) mechanism; that is, conditional upon the covariates in the analysis model and the observed outcomes of the variable being analyzed, the probability of missingness does still depend on the unobserved outcomes of the variable being analyzed.^5^ A third mechanism is missing completely at random (MCAR) in which the observed data are not related to the probability of dropout (after considering covariates).^5^

In practice, MAR is often confused with MCAR. For example, the validity of MAR is often discounted because it is not reasonable to assume participants with missing data would have values similar to participants with no missing data. However, those conditions are what is required for MCAR. In MAR, the predictive distributions are conditioned on covariates and previous values of the dependent variable. Therefore, if a participant was doing worse than group average while observed, MAR projects that this participant would be worse than group average at endpoint.

### 2.3 Analytic approaches

A common approach in the analysis of longitudinal clinical trial data is to fit baseline values of the dependent variable as a covariate.^5(chapter^ ^9), 26–28^ There is a rich literature on covariate adjustment in clinical trials, including covariate imbalance, covariate adjustment, conditional bias, type I error control, gains in efficiency, dropout related to baseline values, etc. ^29–37^

In mITT approaches, only participants with a baseline and at least one post-baseline observation are included. That is, unlike an ITT analysis, mITT does not include all randomized participants. In mITT analyses of the example one data, participants 10 and 20 are not included and the model detailed in Code Fragment 1 is commonly fit to the data. This analysis is referred to as mITTANCOVA because it fits baseline as a covariate in the mITT dataset. An ITT analysis of the same data can be implemented by not excluding participants 10 and 20 from the analysis (ITTANCOVA), thereby complying with the desire to include all randomized participants, and fitting the same model as for the mITT analysis.

A simple and often plausible approach that explicitly addresses the unique nature of participants with no post-baseline data is to impute the first post-baseline assessment as change = 0, but leave all subsequent assessments as missing. Given a short duration of treatment, assuming zero change is defensible in many scenarios. This approach is referred to as ITT_0, or in longer terms, baseline observation carried forward one visit for participants with no post-baseline data.

## Code Fragment 1. SAS and R code for fitting baseline severity as a covariate in MMRM – the ITT_ANCOVA, mITT_ANCOVA, and ITT_0 Analyses

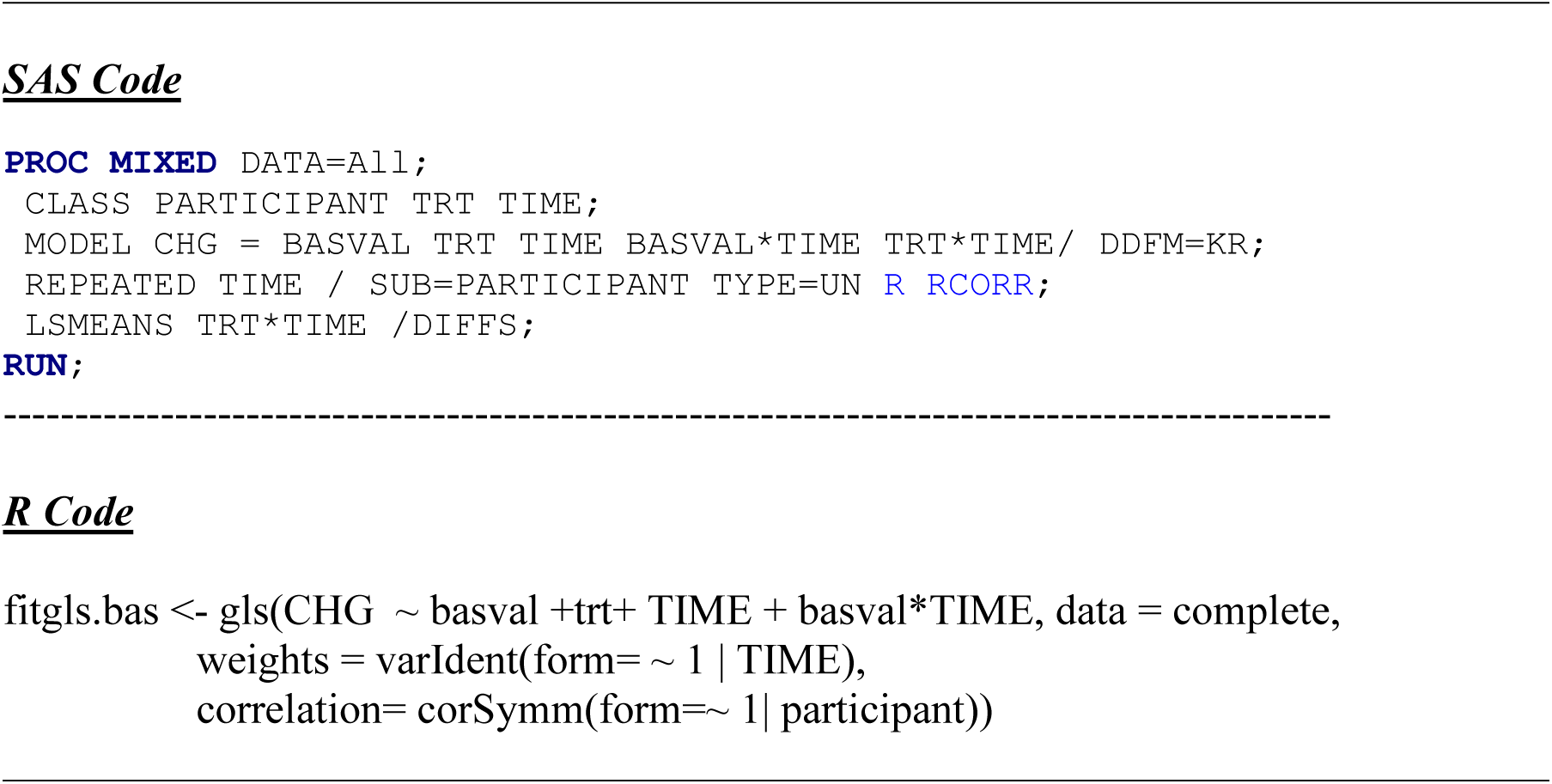

In addition to considering baseline values as a fixed-effect covariate, baseline responses can also be a random effect when included as part of the response vector – the responses at Time 0. ^5(chapter^ ^9), 26–28^ Two methods that include baseline as a response are the LDA (longitudinal data analysis) and cLDA (constrained longitudinal data analyses) approaches.^5(chapter^ ^9), 26–28^ The data input for analyses for two subjects in the example data set are summarized in Table 3.

**Table 3.**
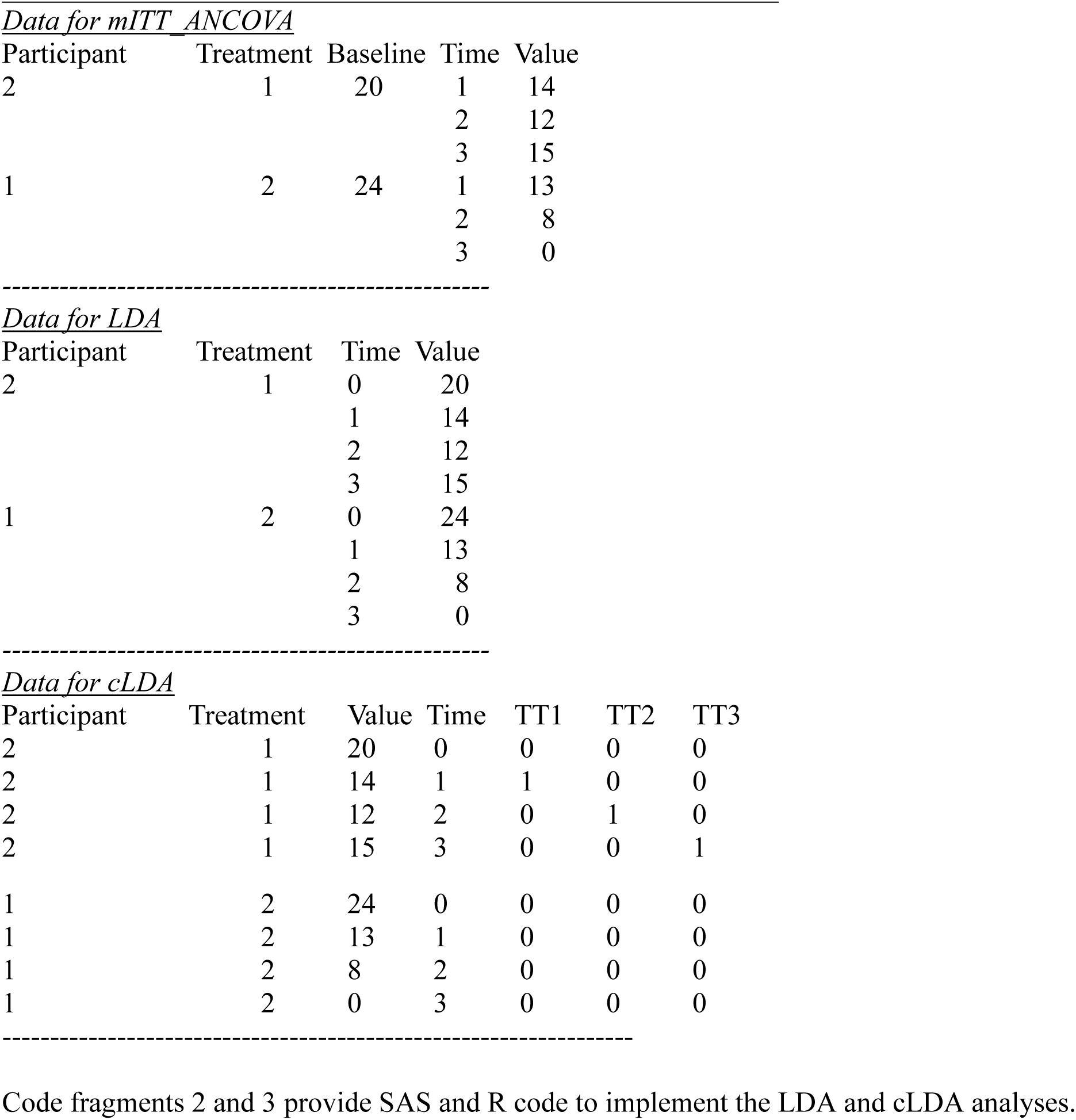
Data for mITT_ANCOVA, LDA, and cLDA analyses

| Participant | Treatment | Baseline | Time | Value |
| --- | --- | --- | --- | --- |
| 2 | 1 | 20 | 1 | 14 |
|  |  |  | 2 | 12 |
|  |  |  | 3 | 15 |
| 1 | 2 | 24 | 1 | 13 |
|  |  |  | 2 | 8 |
|  |  |  | 3 | 0 |

Data for LDA
| Participant | Treatment | Time | Value |
| --- | --- | --- | --- |
| 2 | 1 | 0 | 20 |
|  |  | 1 | 14 |
|  |  | 2 | 12 |
|  |  | 3 | 15 |
| 1 | 2 | 0 | 24 |
|  |  | 1 | 13 |
|  |  | 2 | 8 |
|  |  | 3 | 0 |

Data for cLDA
| Participant | Treatment | Value | Time | TT1 | TT2 | TT3 |
| --- | --- | --- | --- | --- | --- | --- |
| 2 | 1 | 20 | 0 | 0 | 0 | 0 |
| 2 | 1 | 14 | 1 | 1 | 0 | 0 |
| 2 | 1 | 12 | 2 | 0 | 1 | 0 |
| 2 | 1 | 15 | 3 | 0 | 0 | 1 |
| 1 | 2 | 24 | 0 | 0 | 0 | 0 |
| 1 | 2 | 13 | 1 | 0 | 0 | 0 |
| 1 | 2 | 8 | 2 | 0 | 0 | 0 |
| 1 | 2 | 0 | 3 | 0 | 0 | 0 |

For the LDA and cLDA analyses, data are arranged so that there are four responses per participant, with baseline being the Time = 0 response and post-baseline observations at times 1, 2, and 3. For the cLDA analysis additional steps are required to create the time specific treatment indicators TT1-TT3. These dummy variables are components of the treatment-by-time interaction: TTk=I(TIMEk*TRT) for k=1,2,3, where I(.) is the indicator function, returning 1 when the argument is TRUE and 0 if FALSE. ^27,28^

The constraint in cLDA is that the estimated means of baseline values are equal in the intervention groups, which is reasonable under randomization. The TT0 variable is not fit in the model in order to constrain lsmeans by omitting the (Time=0)*Trt interaction term.^27,28^

## Code Fragment 2. SAS and R code for fitting an LDA model

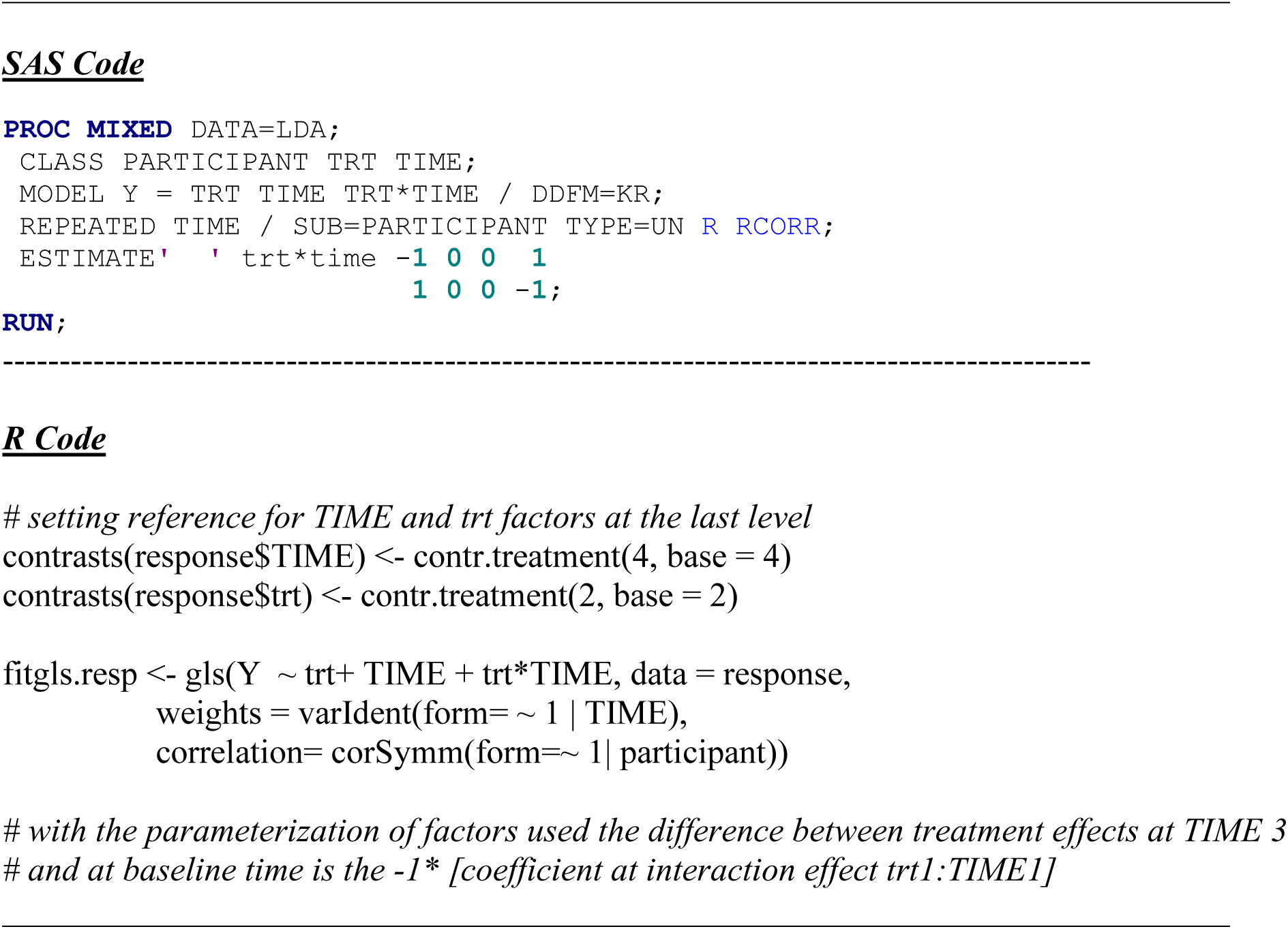

## Code Fragment 3. SAS and R code for fitting a cLDA model

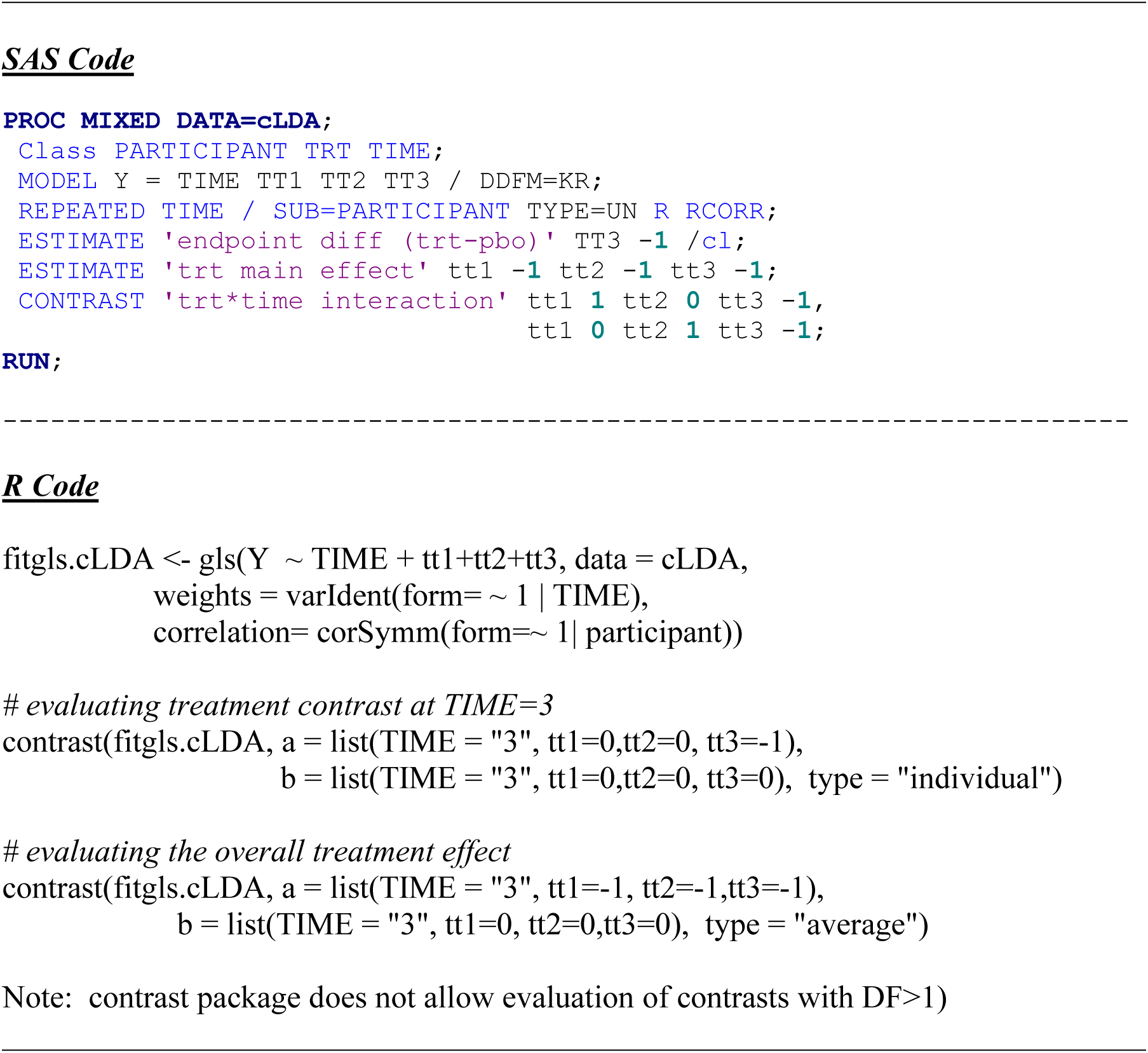

Multiple imputation (MI) is another approach to dealing with missing data from participants that have no post baseline data. ^38–43^ Although a number of implementations are possible, the same imputation model that is used for partially complete sequences can also be used for participants with no post-baseline data. No post-baseline data can be viewed as one of the possible monotone missing data sequences.

Code Fragment 4 provides the SAS and R code for monotone missing data patterns that impute the missing values for participants that have 1, 2, or all 3 post-baseline observations missing. After imputation, the multiple imputed data sets can be analyzed with any of the aforementioned longitudinal analyses with results combined using Rubin’s rules.^40^ In this paper, when MI is used data are analyzed with the ITTANCOVA approach specified above. Should non-monotone missingness occur, several approaches can be considered. For example, MCMC can be used to first impute the intermittent missing values, followed by imputation of the remaining monotone missing values^5^, or a fully conditional specification can be used.^42, 43^

## Code Fragment 4. SAS Code for multiple imputation of missing post-baseline data

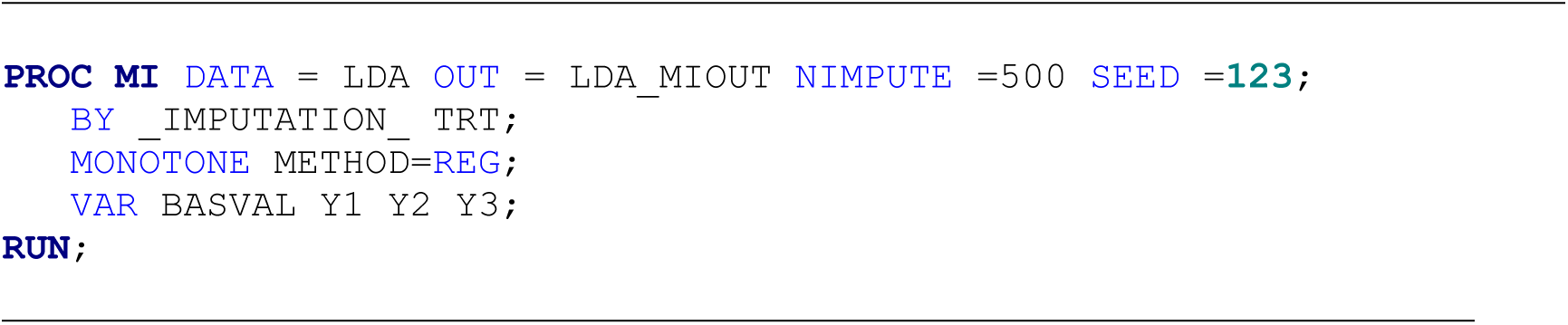

All of the methods mentioned thus far are based on assuming the missing data arise from a missing at random mechanism in targeting a hypothetical strategy of dealing with ICEs to estimate what would have been observed if the ICEs had not occurred. However, MI could also be used to implement MNAR models, either as a primary analysis for relevant estimands or as a sensitivity analysis to the aforementioned MAR-based approaches. For example, the return to baseline approach could be used; if return to baseline is not a reasonable assumption, the reference-based family of imputations can be used to impute missing values by making qualitative reference to another arm in the trial. ^5(chapter^ ^18), 21–25^

In reference-based imputation, deviations from MAR are created by assuming that imputed values take on the characteristics of a reference group, which can be accomplished by imputing missing values for the active arm and the placebo arm based on a model developed from the placebo arm. ^5(chapter^ ^18), 21–25^

Imputing placebo-like outcomes is often a plausibly conservative departure from MAR based on the assumption that if participants do not take the drug, they will not benefit from it. Code fragment 5 provides the SAS code to implement the copy-reference multiple imputation approach. After imputation, the multiple imputed data sets can be analyzed with any of the aforementioned longitudinal analyses with results combined using Rubin’s rules. ^22,23,40^

## Code Fragment 5. SAS Code for the copy reference method of reference-based imputation

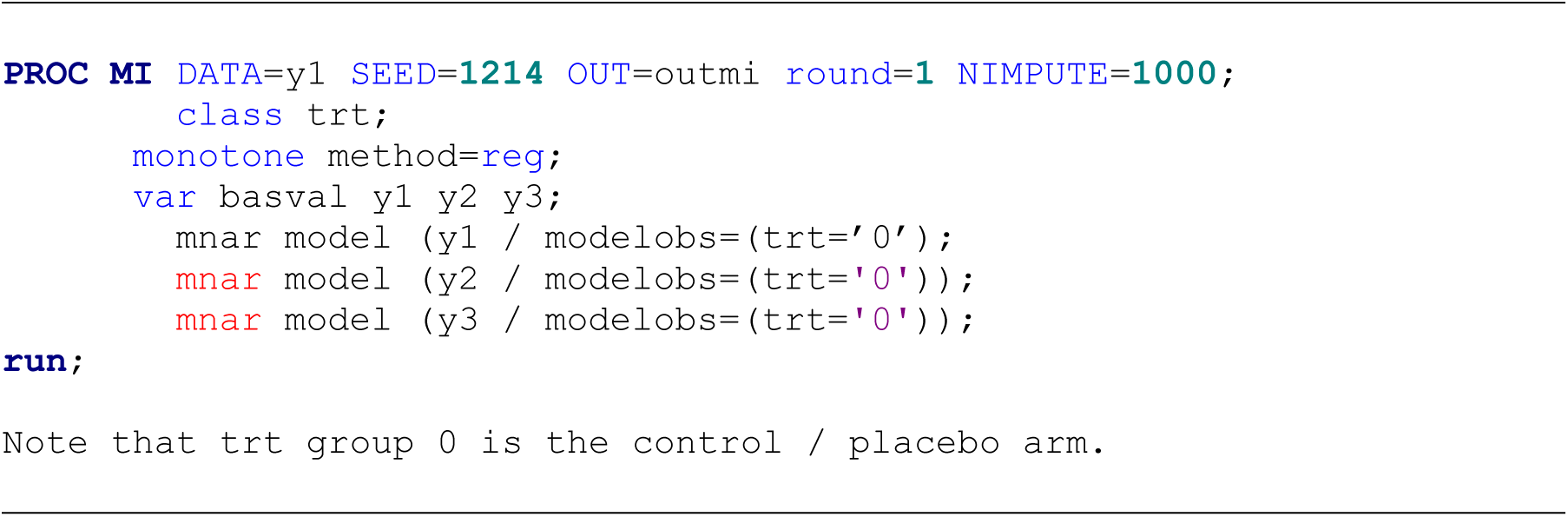

When there are no missing data, the point estimates of the treatment contrasts are identical for mITTANCOVA, ITTANCOVA, and cLDA. The variance estimates, however, can differ because the variance estimate from the ANCOVA model is a conditional variance.^19^ With missing data, these methods may show small differences, typically favoring cLDA.^27^ In scenarios with non-normal data, cLDA provided better confidence interval coverage, power, and Type I error control than the ANCOVA methods.^27^ It is also well known that MI and direct likelihood methods, when implemented in a similar manner, provide asymptotically similar estimates as the size of the data set and number of imputations increases. ^5^ ^39, 42^

Some additional subtle differences between the analytic methods discussed here include the following. Whenever baseline severity is included as a covariate, tests for the treatment main effect and treatment-by-time interaction are more meaningful than in the LDA or cLDA approaches. Treatment main effects in LDA and cLDA are diluted because at Time 0 (baseline) there is no difference between treatments. This zero difference at Time 0 can also inflate the treatment-by-time interaction term in LDA and cLDA.

If baseline severity is considered a fixed effect, then the ANCOVA models are appropriate. The ANCOVA models address conditional estimands – the treatment effect at the mean baseline value. If baseline severity is considered a random effect, then the LDA and cLDA models are appropriate. By considering baseline severity as a random effect, the estimand is not conditional on the current baseline values, inference can be extended to the general population of participants from which the sample was drawn.

### 2.4 Results from the example one data set

Data from the example one data set were analyzed using the models as specified in the code fragments. Key aspects of the results from the various MAR-based methods of accounting for participants with no post-baseline data in the example one data set are summarized in Table 4. Results are identical for the ITTANCOVA and mITTANCOVA because in the ITTANCOVA the two participants with a baseline but no post-baseline data do not contribute information to the estimation of post-baseline parameters.

**Table 4.** Summary of Results from Example One

|  | ITT | mITT | LDA | cLDA | MI | ITT_0 | CR |
| --- | --- | --- | --- | --- | --- | --- | --- |
| # Participants | 20 | 18 | 20 | 20 | 20 | 20 | 20 |
| # Obs Read | 60 | 54 | 80 | 80 | 80 | 60 | 80 |
| # Obs Used | 54 | 54 | 74 | 74 | 74 | 56 | 74 |
| -2 Res Log Likelihood | 267.1 | 267.1 | 367.3 | 370.4 | NA | 284.4 | NA |
| Res variance Time 3 | 14.65 | 14.65 | 14.69 | 14.65 | NA | -15.12 | NA |
| LSMEAN trt 1 Time 3 | -11.51 | -11.51 | -12.11 | -11.67 | -11.52 | -11.36 | -11.52 |
| LSMEAN trt 2 Time 3 | -16.26 | -16.26 | -16.27 | -16.42 | -16.51 | -16.16 | -16.04 |
| LSMEAN DIFF Time 3 | 4.75 | 4.75 | 4.15 | 4.75 | 4.99 | 4.80 | 4.53 |
| SE LSMEAN DIFF Time 3 | 1.80 | 1.80 | 2.15 | 1.86 | 2.06 | 1.84 | 2.24 |
| LSMEAN DIFF p-value | 0.0189 | 0.0189 | 0.0714 | 0.0212 | 0.0152 | 0.0200 | 0.0435 |

The treatment contrast from cLDA, mITT– and ITTANCOVA are identical, as is the residual variance at Time 3. Minor differences in degrees-of-freedom and in the correlations between the repeated measures resulted in a slightly larger standard error and p-value for cLDA.

Results for ITT_0, and to a somewhat lesser degree for MI, were also similar to cLDA and the ANCOVA methods, with a MI having a slightly larger standard error. The LDA results stand furthest apart, with a smaller estimated treatment difference and a larger standard error of the treatment contrast, leading to a greater p-value compared with the methods that constrain baseline values. In the example data the mean baseline values differed by 0.80, which explains most of the difference in results between the LDA and other methods. As expected, results from the MNAR-based CR show an attenuated treatment effect and larger standard error compared to the MAR-based methods.

It is important to consider the results of the example data set as a contrived example to make it easier to see differences between the analyses. For example, data from the subject with the greatest baseline value had no post-baseline data, a feature implemented to create differences between the various ANCOVA approaches and LDA.

### 2.5. Results from example two data sets

An important difference in the analyses of the example two data sets compared with example one is that the example two analyses included the fixed effect of investigative site. All other aspects of the models were the same. Results from the various methods of accounting for participants with no post-baseline data in the example two data sets are summarized in Table 5.

**Table 5.** Summary of Results from Example Two

| Method | Low Dropout Data set |  |  | High Dropout Dataset |  |  |
| --- | --- | --- | --- | --- | --- | --- |
|  | LSMEAN | StdErr | Probt | LSMEAN | STdErr | Probt |
| ITT | 1.8567 | 0.6989 | 0.0086 | 2.3710 | 1.0304 | 0.0230 |
| mITT | 1.8567 | 0.6989 | 0.0086 | 2.3710 | 1.0304 | 0.0230 |
| LDA | 2.2144 | 0.7705 | 0.0045 | 2.8835 | 1.1164 | 0.0108 |
| cLDA | 1.8320 | 0.7016 | 0.0098 | 2.4722 | 1.1010 | 0.0263 |
| MI | 1.8591 | 0.7059 | 0.0085 | 2.4593 | 1.1191 | 0.0280 |
| ITT_0 | 1.8731 | 0.6988 | 0.0080 | 2.3402 | 1.0579 | 0.0287 |
| CR | 1.7251 | 0.7075 | 0.0148 | 1.6827 | 1.0624 | 0.1133 |

Results are again identical for ITTANCOVA and mITTANCOVA, and results from ITT_0 and MI are similar to the ANCOVA results. Unlike in the example one dataset, the cLDA results differ from the ITTANCOVA and mITTANCOVA results. This was due to the example two data including the fixed effect of site, that was not included in the example one analyses.

Including baseline values in the response vector provide additional data used in estimating the site effects, which in turn influenced estimates of other effects in the model. Greater differences between methods are expected as data become more unbalanced.

The LDA results were again notably differen. This disparity in results stems from LDA being the only method that does not in some way constraint baseline values to be equal for the two treatments. In the raw data, the baseline means differed by approximately 0.5 points in the low dropout dataset and 1.4 points in the high dropout dataset.

Among the “constrained” methods (ITTANCOVA, mITTANCOVA, cLDA, ITT_O, and MI) results varied by 0.0411 (1.8320 to 1.8731) in the low dropout dataset. In the high dropout dataset, results among the constrained methods varied by 0.1320 which is ∼3-fold greater than in the low dropout dataset. Of course, greater sensitivity to missing data is expected when the rate of missing data is higher. Despite the larger point estimates from MAR methods in the high dropout dataset, the treatment contrast from CR was greater in the low dropout dataset because the attenuation of the treatment effect in CR is roughly proportional to the percent of missing data in the active arm.

## 3 Simulation Study

### 3.1. Data generation and design

To further understand various approaches to handling missing data from participants with no post-baseline data we conducted a simulation study. The primary objective of the simulation study was to compare results from the analyses detailed in Section 2 across several scenarios and missing data mechanisms. The model used to simulate the data was as follows:

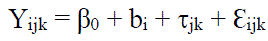

where:

β_0_ is the fixed intercept

b_i_ is the random effect of the ith participant, i = 1,…, 200 b_i_ ∼ N(0,σ^2^_participant_)

τ_jk_ are the treatment by time interaction and main treatment effects for treatment j at time k. j=1 (control) and 2 (active); k = 0 (baseline) and k = 1-3 (times 1-3). For identifiability, τ_1k_=0, for all times (k) and τ_j0_=0 for all treatments (j).

Ɛ_ijk_ is the random residual effect for participant i in treatment j at time k. Ɛ_ijk_ ∼ N(0, σ^2^_Ɛ_).

Four scenarios were simulated as a 2 x 2 factorial, with 2 levels of correlation between baseline and post-baseline values: 0.25 (low) or 0.75 (high) 2 levels of treatment effect: 0.0 or 0.4 standardized treatment effect (Cohen’s D)

The variances of random effects in the low correlation scenarios were σ^2^_participant_=0.25, σ^2^_Ɛ_ =0.75 The variances of random effects in the high correlation scenarios were σ^2^_participant_=0.75, σ^2^_Ɛ_ =0.25 In all scenarios, outcomes were normalized to have the standard deviation in Y = 1.0.

The true treatment-by-time mean profiles in the null hypothesis scenarios of 0.0 treatment effect were constant and equal 10.0 for both treatments at all times. The true treatment-by-time means, β_0_ + τ_jk_ (j=1,2; k=0,..,3), for the alternative hypothesis scenarios 0.4 treatment effect were:

|  | Time | 0 | 1 | 2 | 3 |
| --- | --- | --- | --- | --- | --- |
| Treatment 1 |  | 10.0 | 10.0 | 10.0 | 10.0 |
| Treatment 2 |  | 10.0 | 9.8 | 9.7 | 9.6 |

Each data set included 100 participants per arm and 5000 data sets were simulated for each scenario. The following four missing data mechanisms were applied to each simulated data set to delete all post-baseline data for ∼10% of participants. In addition, in each data set another 10% of the participants had missing data at Visit 3 as a result of a completely random mechanism.

|  |  |  |
| --- | --- | --- |
| **Random** | (Random) | 10% probability of participants having all post-baseline values deleted randomly, unrelated to treatment or outcome |
| **Treatment related** | (TRT_REL) | 20% probability of participants in treatment 1 having all post-baseline values deleted, unrelated to outcome |
| **Outcome related** | (OUT_REL) | 20% probability of participants with outcomes at the first post-baseline time that are above 10.0 having all post-baseline data deleted, same mechanism in each treatment |
| **Treatment & Outcome related** | (TRTOUT_REL) | 40% probability of participants in group 2 with outcomes at the first post-baseline time that are > 10.0 having all post-baseline data deleted |

In scenarios with outcome-related missingness, the missing data mechanisms was MNAR because with no observed data on these subjects, missingness was not explained by the observed data – it depended on the unobserved data. The missing data mechanism for the random scenarios was MCAR, and the mechanism for the treatment related scenarios was covariate-dependent MCAR.

A total of 80,000 data sets were analyzed: 5000 data sets for each of 4 missing data mechanisms in each of 4 scenarios (correlation high or low, treatment effect 0. or 0.4.

### 3.3 Analyses

Each data set was analyzed using the methods and models described in Section 2; namely, mITTANCOVA, ITTANCOVA, LDA, cLDA, MI, ITT_0, and CR. The mean treatment contrasts, mean standard errors of the treatment contrasts, and mean treatment contrast p-value were summarized for each analysis method, along with the % of treatment contrasts that were statistically significant (power / Type I error). The standard deviation in treatment contrasts was also summarized and compared with the mean standard error to assess whether the standard errors accurately reflected the uncertainty in the results.

### 3.4 Results

Descriptive statistics for the various simulation scenarios and missing data mechanisms are summarized in Table 6. With 10% of data missing completely at random in every scenario, the percent of subjects with no post-baseline data is the difference between the observed completion percentage and 90%.

**Table 6.** Descriptive statistics for simulated data.

| **Mechanism** | **% complete data at Visit 3** |  | **Means at Visit 3** |  | **Observed mean** |
| --- | --- | --- | --- | --- | --- |
|  | **Group 1** | **Group 2** | **Group 1** | **Group 2** | **Difference** |
| *Delta = 0.4, Correlation = strong* |  |  |  |  |  |
| RANDOM | 80.9 | 81.1 | 10.0034 | 9.5963 | 0.4068 |
| TRT_REL | 89.9 | 72.1 | 10.0028 | 9.5958 | 0.4070 |
| OUT_REL | 80.9 | 82.4 | 9.9366 | 9.5326 | 0.4040 |
| TRTOUT_REL | 89.9 | 69.2 | 10.0004 | 9.7495 | 0.2509 |
| *Delta = 0.0, Correlation = weak* |  |  |  |  |  |
| RANDOM | 81.0 | 81.1 | 10.0034 | 9.9963 | 0.0071 |
| TRT_REL | 89.8 | 72.1 | 10.0028 | 9.9958 | 0.0070 |
| OUT_REL | 80.6 | 81.0 | 9.9343 | 9.9302 | 0.0041 |
| TRTOUT_REL | 89.9 | 69.2 | 10.0028 | 10.1461 | -0.1433 |
| *Delta = 0.4, Correlation = weak* |  |  |  |  |  |
| RANDOM | 80.9 | 81.1 | 10.0036 | 9.5967 | 0.4069 |
| TRT_REL | 89.9 | 72.1 | 10.0031 | 9.5964 | 0.4065 |
| OUT_REL | 80.8 | 82.5 | 9.9806 | 9.5776 | 0.4030 |
| TRTOUT_REL | 89.9 | 69.2 | 10.0031 | 9.6479 | 0.3552 |
| *Delta = 0.0, Correlation = weak* |  |  |  |  |  |
| RANDOM | 80.9 | 81.1 | 10.0036 | 9.9967 | 0.0069 |
| TRT_REL | 89.9 | 72.1 | 10.0031 | 9.9967 | 0.0064 |
| OUT_REL | 80.9 | 81.1 | 9.9806 | 9.9749 | 0.0057 |
| TRTOUT_REL | 89.9 | 72.0 | 10.0031 | 10.0471 | -0.0440 |

With a random missingness mechanism, % completion was similar across groups at ∼80%. With treatment related missingness, % completion was ∼90% for group 1 (control) and 72% for Group 2. With outcome related missingness, % completion was similar across groups in the scenarios with no treatment effect, but as expected, was slightly higher in Group 2 in scenarios with a treatment effect favoring group 2. With treatment and outcome related missingness, Group 1 had 90% completion and Group 2 had 69% completion.

The random, treatment-related, and outcome-related missing data mechanisms did not bias treatment group differences. Although the within group means were biased by outcome related dropout, this bias was equal in the two treatment arms when there was no treatment effect and nearly equal when there was a treatment effect. Only when missingness was treatment and outcome related was appreciable bias seen in the observed data. The greatest bias was seen with strong correlation between repeated measurements. The maximum bias was ∼ 0.15, which in relative terms was 3/8 (37.5%) the magnitude of the true treatment effect.

Results from the various analyses are summarized for the 4 simulation scenarios in Tables 7a – 7d. Results for the MAR-based methods are described first, followed by results for CR.

As in the example data sets, treatment contrasts from ITTANCOVA, mITTANCOVA, and cLDA were equal, and mean standard error and mean p-values were also very similar across these 3 methods. The mean treatment contrasts from MI were similar to the ANCOVAs and cLDA; however, the mean standard error and mean p-values were greater and % significant slightly lower. This result was expected given the well-known inflation of variance in MI ^17,31,33^

For ITT_0, the mean treatment contrasts, standard errors, p-values and % significant were all similar to the ANCOVAs and cLDA.

Results from LDA showed mean treatment contrasts similar to other methods, but with larger mean standard errors and p values, leading to lower % significant. The difference in % significant was more pronounced in the scenarios with weak correlation where power was lower due to greater error variance. In these scenarios, LDA had ∼15% lower power and MI had about 3% lower power than the other methods.

The only mechanism that led to bias was treatment and outcome related missingness. In these scenarios the mean treatment contrasts underestimated the true treatment difference of 0.40 by ∼0.035 for all MAR-based methods. As a percentage of the true treatment effect, bias was about 9%, which is considerably smaller than the 37.5% relative bias in the observed data. The highest Type I error rate in any scenario was 6.4%. The highest Type I error rate for MI was 5.5% and the highest Type I error rate for ITT_0 was 5.1.

For CR, mean treatment contrasts in scenarios with a true effect ranged from 0.25 – 0.35, attenuating the treatment effect approximately 10% to 40% compared with other methods, with greater mean SE and lower power. However, given CR can be used as either a sensitivity analysis to the MAR-based methods, or as a primary means of addressing other hypothetical estimands that assume for some or all of the ICEs, that participants with the relevant ICEs have placebo-like outcomes, the attenuated treatment effect is not bias. In Scenarios with no treatment effect the Type I error rate from CR was well below the nominal rate. With the CR analyses, the mean standard error was appreciably greater than the standard deviation in treatment contrasts, consistent with the inflation of variance in the reference-based family of methods.^36, 37^

**Table 7a.** Results from Various Analysis Methods in Simulated Data – delta = 0.40, Correlation = strong.

| **Missingness mechanism = Random** |  |  |  |  |  |
| --- | --- | --- | --- | --- | --- |
| Method | Mean Contrast | Std Dev Contrast | Mean SE Contrast | Mean Pval Contrast | % Sig Contrast |
| ITTANCOVA | 0.4034 | 0.1042 | 0.1030 | 0.0056 | 97.6 |
| mITTANCOVA | 0.4034 | 0.1042 | 0.1030 | 0.0056 | 97.6 |
| cLDA | 0.4034 | 0.1042 | 0.1030 | 0.0056 | 97.6 |
| LDA | 0.4030 | 0.1101 | 0.1087 | 0.0089 | 95.8 |
| MI | 0.4033 | 0.1056 | 0.1086 | 0.0077 | 96.7 |
| ITT_0 | 0.3947 | 0.1027 | 0.1015 | 0.0060 | 97.3 |
| CR | 0.3404 | 0.0884 | 0.1042 | 0.0123 | 93.9 |
| **Missingness mechanism = Treatment Related** |  |  |  |  |  |
| ITTANCOVA | 0.4034 | 0.1041 | 0.1036 | 0.0059 | 97.7 |
| mITTANCOVA | 0.4034 | 0.1041 | 0.1036 | 0.0059 | 97.7 |
| cLDA | 0.4034 | 0.1041 | 0.1036 | 0.0059 | 97.6 |
| LDA | 0.4030 | 0.1099 | 0.1093 | 0.0090 | 95.7 |
| MI | 0.4034 | 0.1053 | 0.1093 | 0.0079 | 96.2 |
| ITT_0 | 0.3863 | 0.1027 | 0.1020 | 0.0074 | 97.0 |
| CR | 0.3025 | 0.0794 | 0.1051 | 0.0216 | 88.4 |
| **Missingness mechanism = Outcome Related** |  |  |  |  |  |
| ITTANCOVA | 0.4027 | 0.1033 | 0.1025 | 0.0054 | 97.7 |
| mITTANCOVA | 0.4027 | 0.1033 | 0.1025 | 0.0054 | 97.7 |
| cLDA | 0.4027 | 0.1033 | 0.1025 | 0.0054 | 97.7 |
| LDA | 0.4023 | 0.1092 | 0.1083 | 0.0088 | 96.0 |
| MI | 0.4023 | 0.1091 | 0.1083 | 0.0087 | 96.1 |
| ITT_0 | 0.3958 | 0.1020 | 0.1012 | 0.0056 | 97.6 |
| CR | 0.3456 | 0.0889 | 0.1035 | 0.0109 | 94.9 |
| **Missingness mechanism = Treatment & Outcome Related** |  |  |  |  |  |
| ITTANCOVA | 0.3641 | 0.1052 | 0.1051 | 0.0144 | 93.6 |
| mITTANCOVA | 0.3641 | 0.1052 | 0.1051 | 0.0144 | 93.6 |
| cLDA | 0.3641 | 0.1052 | 0.1048 | 0.0142 | 93.5 |
| LDA | 0.3637 | 0.1109 | 0.1104 | 0.0203 | 90.4 |
| MI | 0.3639 | 0.1075 | 0.1117 | 0.0196 | 90.6 |
| ITT_0 | 0.3655 | 0.1037 | 0.1030 | 0.0123 | 94.3 |
| CR | 0.2603 | 0.0775 | 0.1064 | 0.0478 | 74.0 |

**Table 7b.**
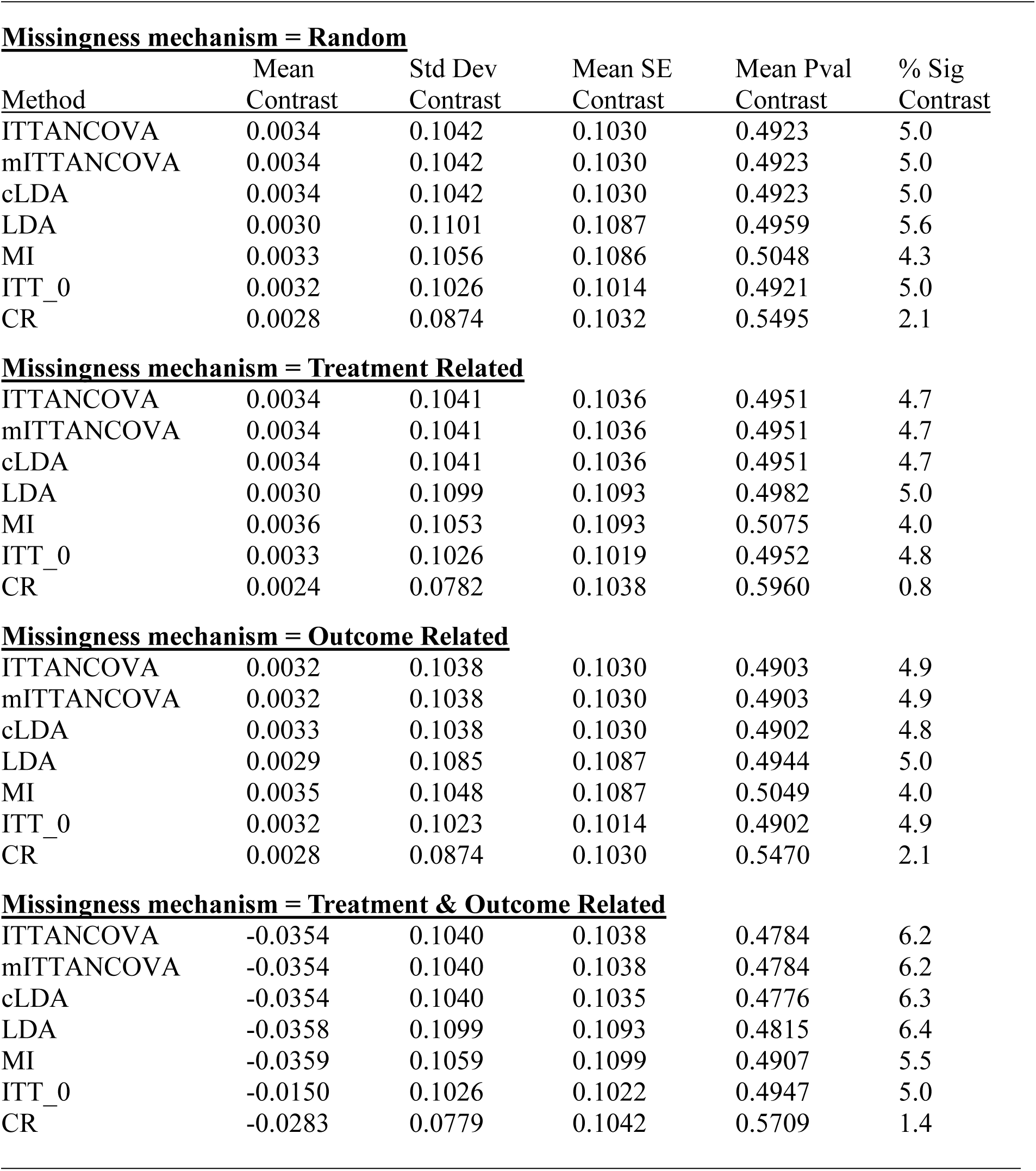
Results from Various Analysis Methods in Simulated Data – Delta = 0.0, Correlation = strong.

| **Missingness mechanism = Random** |  |  |  |  |  |
| --- | --- | --- | --- | --- | --- |
| Method | Mean Contrast | Std Dev Contrast | Mean SE Contrast | Mean Pval Contrast | % Sig Contrast |
| ITTANCOVA | 0.0034 | 0.1042 | 0.1030 | 0.4923 | 5.0 |
| mITTANCOVA | 0.0034 | 0.1042 | 0.1030 | 0.4923 | 5.0 |
| cLDA | 0.0034 | 0.1042 | 0.1030 | 0.4923 | 5.0 |
| LDA | 0.0030 | 0.1101 | 0.1087 | 0.4959 | 5.6 |
| MI | 0.0033 | 0.1056 | 0.1086 | 0.5048 | 4.3 |
| ITT_0 | 0.0032 | 0.1026 | 0.1014 | 0.4921 | 5.0 |
| CR | 0.0028 | 0.0874 | 0.1032 | 0.5495 | 2.1 |
| **Missingness mechanism = Treatment Related** |  |  |  |  |  |
| ITTANCOVA | 0.0034 | 0.1041 | 0.1036 | 0.4951 | 4.7 |
| mITTANCOVA | 0.0034 | 0.1041 | 0.1036 | 0.4951 | 4.7 |
| cLDA | 0.0034 | 0.1041 | 0.1036 | 0.4951 | 4.7 |
| LDA | 0.0030 | 0.1099 | 0.1093 | 0.4982 | 5.0 |
| MI | 0.0036 | 0.1053 | 0.1093 | 0.5075 | 4.0 |
| ITT_0 | 0.0033 | 0.1026 | 0.1019 | 0.4952 | 4.8 |
| CR | 0.0024 | 0.0782 | 0.1038 | 0.5960 | 0.8 |
| **Missingness mechanism = Outcome Related** |  |  |  |  |  |
| ITTANCOVA | 0.0032 | 0.1038 | 0.1030 | 0.4903 | 4.9 |
| mITTANCOVA | 0.0032 | 0.1038 | 0.1030 | 0.4903 | 4.9 |
| cLDA | 0.0033 | 0.1038 | 0.1030 | 0.4902 | 4.8 |
| LDA | 0.0029 | 0.1085 | 0.1087 | 0.4944 | 5.0 |
| MI | 0.0035 | 0.1048 | 0.1087 | 0.5049 | 4.0 |
| ITT_0 | 0.0032 | 0.1023 | 0.1014 | 0.4902 | 4.9 |
| CR | 0.0028 | 0.0874 | 0.1030 | 0.5470 | 2.1 |
| **Missingness mechanism = Treatment & Outcome Related** |  |  |  |  |  |
| ITTANCOVA | -0.0354 | 0.1040 | 0.1038 | 0.4784 | 6.2 |
| mITTANCOVA | -0.0354 | 0.1040 | 0.1038 | 0.4784 | 6.2 |
| cLDA | -0.0354 | 0.1040 | 0.1035 | 0.4776 | 6.3 |
| LDA | -0.0358 | 0.1099 | 0.1093 | 0.4815 | 6.4 |
| MI | -0.0359 | 0.1059 | 0.1099 | 0.4907 | 5.5 |
| ITT_0 | -0.0150 | 0.1026 | 0.1022 | 0.4947 | 5.0 |
| CR | -0.0283 | 0.0779 | 0.1042 | 0.5709 | 1.4 |

**Table 7c.** Results from Various Analysis Methods in Simulated Data – Delta = 0.40, Correlation = weak.

| **Missingness mechanism = Random** |  |  |  |  |  |
| --- | --- | --- | --- | --- | --- |
| Method | Mean Contrast | Std Dev Contrast | Mean SE Contrast | Mean Pval Contrast | % Sig Contrast |
| ITTANCOVA | 0.4062 | 0.1547 | 0.1523 | 0.0589 | 74.3 |
| mITTANCOVA | 0.4062 | 0.1547 | 0.1523 | 0.0589 | 74.3 |
| cLDA | 0.4062 | 0.1547 | 0.1523 | 0.0589 | 74.3 |
| LDA | 0.4054 | 0.1879 | 0.1853 | 0.1175 | 57.7 |
| MI | 0.4058 | 0.1568 | 0.1577 | 0.0654 | 71.5 |
| ITT_0 | 0.4020 | 0.1544 | 0.1519 | 0.0608 | 73.5 |
| CR | 0.3364 | 0.1283 | 0.1533 | 0.0905 | 60.3 |
| **Missingness mechanism = Treatment Related** |  |  |  |  |  |
| ITTANCOVA | 0.4060 | 0.1545 | 0.1532 | 0.0597 | 73.6 |
| mITTANCOVA | 0.4060 | 0.1545 | 0.1532 | 0.0597 | 73.6 |
| cLDA | 0.4060 | 0.1545 | 0.1532 | 0.0597 | 73.6 |
| LDA | 0.4052 | 0.1871 | 0.1861 | 0.1179 | 57.3 |
| MI | 0.4060 | 0.1564 | 0.1590 | 0.0663 | 70.8 |
| ITT_0 | 0.3980 | 0.1543 | 0.1528 | 0.0641 | 72.4 |
| CR | 0.2987 | 0.1150 | 0.1543 | 0.1187 | 48.4 |
| **Missingness mechanism = Outcome Related** |  |  |  |  |  |
| ITTANCOVA | 0.4047 | 0.1534 | 0.1516 | 0.0583 | 74.4 |
| mITTANCOVA | 0.4047 | 0.1534 | 0.1516 | 0.0583 | 74.4 |
| cLDA | 0.4047 | 0.1534 | 0.1516 | 0.0583 | 74.4 |
| LDA | 0.4040 | 0.1864 | 0.1848 | 0.1163 | 58.0 |
| MI | 0.4051 | 0.1548 | 0.1569 | 0.0637 | 71.7 |
| ITT_0 | 0.4017 | 0.1530 | 0.1513 | 0.0597 | 73.9 |
| CR | 0.3410 | 0.1298 | 0.1525 | 0.0866 | 61.4 |
| **Missingness mechanism = Treatment & Outcome Related** |  |  |  |  |  |
| ITTANCOVA | 0.3670 | 0.1552 | 0.1550 | 0.0907 | 65.5 |
| mITTANCOVA | 0.3670 | 0.1552 | 0.1550 | 0.0907 | 65.5 |
| cLDA | 0.3671 | 0.1553 | 0.1549 | 0.0906 | 65.6 |
| LDA | 0.3663 | 0.1877 | 0.1876 | 0.1556 | 49.3 |
| MI | 0.3669 | 0.1579 | 0.1615 | 0.1011 | 62.3 |
| ITT_0 | 0.3723 | 0.1552 | 0.1545 | 0.0857 | 66.8 |
| CR | 0.2565 | 0.1115 | 0.1560 | 0.1780 | 32.9 |

**Table 7d.** Results from Various Analysis Methods in Simulated Data – Delta = 0.0, Correlation = weak.

| **Missingness mechanism = Random** |  |  |  |  |  |
| --- | --- | --- | --- | --- | --- |
| Method | Mean Contrast | Std Dev Contrast | Mean SE Contrast | Mean Pval Contrast | % Sig Contrast |
| ITTANCOVA | 0.0061 | 0.1547 | 0.1523 | 0.4927 | 4.9 |
| mITTANCOVA | 0.0061 | 0.1547 | 0.1523 | 0.4927 | 4.9 |
| cLDA | 0.0062 | 0.1547 | 0.1523 | 0.4927 | 4.8 |
| LDA | 0.0054 | 0.1879 | 0.1853 | 0.4995 | 4.7 |
| MI | 0.0058 | 0.1568 | 0.1579 | 0.4995 | 4.7 |
| ITT_0 | 0.0060 | 0.1544 | 0.1519 | 0.4927 | 4.9 |
| CR | 0.0048 | 0.1277 | 0.1526 | 0.5549 | 2.1 |
| **Missingness mechanism = Treatment Related** |  |  |  |  |  |
| ITTANCOVA | 0.0060 | 0.1545 | 0.1532 | 0.4927 | 4.8 |
| mITTANCOVA | 0.0060 | 0.1545 | 0.1532 | 0.4927 | 4.8 |
| cLDA | 0.0060 | 0.1545 | 0.1532 | 0.4927 | 4.8 |
| LDA | 0.0052 | 0.1871 | 0.1861 | 0.4981 | 5.0 |
| MI | 0.0061 | 0.1564 | 0.1588 | 0.4994 | 4.4 |
| ITT_0 | 0.0059 | 0.1543 | 0.1528 | 0.4921 | 4.8 |
| CR | 0.0040 | 0.1139 | 0.1533 | 0.5909 | 1.0 |
| **Missingness mechanism = Outcome Related** |  |  |  |  |  |
| ITTANCOVA | 0.0051 | 0.1540 | 0.1522 | 0.4923 | 5.1 |
| mITTANCOVA | 0.0051 | 0.1540 | 0.1522 | 0.4923 | 5.1 |
| cLDA | 0.0051 | 0.1540 | 0.1522 | 0.4923 | 5.1 |
| LDA | 0.0044 | 0.1867 | 0.1853 | 0.4975 | 5.2 |
| MI | 0.0053 | 0.1561 | 0.1578 | 0.4998 | 4.9 |
| ITT_0 | 0.0051 | 0.1535 | 0.1519 | 0.4925 | 5.0 |
| CR | 0.0041 | 0.1277 | 0.1525 | 0.5545 | 1.7 |
| **Missingness mechanism = Treatment & Outcome Related** |  |  |  |  |  |
| ITTANCOVA | -0.0322 | 0.1567 | 0.1533 | 0.4909 | 5.3 |
| mITTANCOVA | -0.0322 | 0.1567 | 0.1533 | 0.4909 | 5.3 |
| cLDA | -0.0322 | 0.1567 | 0.1532 | 0.4908 | 5.3 |
| LDA | -0.0330 | 0.1866 | 0.1862 | 0.4934 | 5.2 |
| MI | -0.0328 | 0.1559 | 0.1591 | 0.4979 | 4.9 |
| ITT_0 | -0.0190 | 0.1538 | 0.1529 | 0.4938 | 5.1 |
| CR | -0.0265 | 0.1132 | 0.1536 | 0.5862 | 1.1 |

## 4. Discussion

Dealing with the missing data for participants who are randomized to treatment and have a baseline value but all post-baseline data are missing, is a case of particular interest. The challenge is that with no post-baseline data there is no information about the outcome or the intercurrent events that led to missing data.

The mITT approach of including only participants with a baseline and at least one post-baseline value jeopardizes randomization. Not surprisingly, regulators often ask for an ITT-based analysis with specific mention of how the missing data from participants with no post-baseline data will be handled.

Treatment policy, while on treatment, and principal stratification ICE strategies will typically not address the randomization concerns. Composite and hypothetical strategies can address the concern. Composite strategies are well-suited for scenarios where reasonable and meaningful outcomes can be ascribed for participants with relevant ICEs. This may be challenging for participants with no post-baseline data because there is no information about the outcome or the ICE(s) leading to missing data. Hypothetical strategies are useful when wanting to know what outcomes would have been under various hypothetical conditions/scenarios.

In this paper, we illustrated and compared commonly approaches for the hypothetical strategy of estimating what would have been observed if the participants had not dropped out. We also implemented a copy-reference MNAR based approach using the assumption that drug treated participants who dropped out have placebo-like outcomes. These hypothetical approaches were applied to both participants with no post-baseline data and those with partial post-baseline data. The CR results can be used as either a sensitivity analysis to an MAR-based method or as a primary approach for a hypothetical estimand that assumes a specific MNAR trajectory in which missing data for active arm participants is imputed from the model developed from the placebo arm, thereby imputing placebo-like values for active arm participants with missing data.

Results in the example clinical trial data and simulation study were similar among the various MAR-based methods, especially those that constrained baseline values to be equal for the two treatments. In LDA, the unconstrained method, the standard error were greater leading to as much as 15% lower power.

In the simulated data, estimates of the treatment effect were not biased when the mechanism leading to no post-baseline data was random or treatment related. There was minimal bias when the mechanisms was outcome related, but that bias was approximately equal in the two groups leading to estimates of treatment differences that were not biased. It was only when the mechanism was both treatment and outcome related that bias in the treatment contrasts occurred, and that bias was not large. For example, the highest type I error rates were 6.4% in LDA, 6.3% for cLDA, ITTANCOVA, and mITTANCOVA, 5.5% for MI, and 5.1% for ITT_0 in a scenario where all 10% of the participants who had no post-baseline data all came from one treatment group and would have had below group average outcomes at the 1^st^ post-baseline visit if observed. The CR results, which could be used as a sensitivity analysis, or to address a different estimand, showed the expected attenuated treatment effect.

These results should be interpreted in light of several limitations and considerations. The missingness mechanisms tested in the simulation study were not intended to match any particular clinical trial scenario, but rather scenarios were chosen to clearly show differences between methods and scenarios by using extreme mechanisms as benchmarks.

In actual clinical trial data, there may be multiple intercurrent events to be handled. Therefore, hybrid strategies that use different imputation strategies depending on intercurrent event can be considered^44^. For hybrid strategies, the flexibility of imputation-based methods may be beneficial.

As illustrated with the two studies in example two, the best way of dealing with the missing data was to minimize the amount of it. Missing data in general, including participants with no post-baseline data, had a much smaller effect in the low dropout dataset. Therefore, designs should emphasize participant retention^45, 46^ and include scheduled assessments early enough to minimize loss to follow up prior to post-baseline observation.

## 5. Conclusion

Composite and hypothetical strategies can allow for inclusion of participants with no post-baseline data. Given that there is no information about the treatment effect when there is no post-baseline data, it is not surprising that various models yielded similar estimates of the treatment effect. Treatment effect estimates were not biased when the reason for missing all post-baseline data was random, treatment related, or outcome related. Bias only occurred when treatment and outcome related missingness arose from an MNAR mechanism, and then the bias was not large.

Given the idiosyncratic nature of clinical trials, no universally best analytic approach exists for dealing with participants that have a baseline but no post-baseline data. Analysts can choose among these methods to provide an approach tailored to the situation at hand. Study design and conduct should consider means to minimize the percentage of participants with no post-baseline data as part of the overall consideration of minimizing missing data.

## Data Availability

All data produced in the present study are available upon reasonable request to the authors

